# TMEM147 promotes tumor progression by promoting immunosuppression and serves as a prognostic biomarker for lung adenocarcinoma

**DOI:** 10.1101/2025.08.01.25332641

**Authors:** Lei Ding, Yi Ding

**Author notes:** **Correspondence authors at**:Department of Radiology, The First Affiliated Hospital of Heilongjiang University of Traditional Chinese Medicine, Harbin 150040, China. E-mail addresses (Yi Ding).

## Abstract

TMEM147, an ER membrane protein, is linked to lung adenocarcinoma (LUAD) but its role remains unclear. This study combines bioinformatics and experiments to explore TMEM147’s function in LUAD progression. TMEM147 expression was analyzed using TCGA/GEO data and validated in LUAD cells. Survival analysis assessed its prognostic value. GO/KEGG and ssGSEA revealed functional pathways and immune microenvironment interactions. Transcription factor binding predictions and in vitro assays (migration, invasion, proliferation) evaluated TMEM147’s role. TMEM147 was upregulated in LUAD and correlated with poor outcomes. FLI1 was predicted as a transcriptional regulator. TMEM147 influenced immune cell infiltration and was associated with ribonucleoprotein biogenesis and oxidative phosphorylation (OXPHOS) . Silencing TMEM147 reduced cancer cell migration, invasion, and proliferation, suggesting its potential as a biomarker and therapeutic target.

## 1. Introduction

Globally, lung cancer accounts for approximately 11.4% of the 19.3 million new cancer cases diagnosed annually and is responsible for nearly 20% of all cancer-related deaths, making it one of the deadliest malignancies worldwide. Based on histological characteristics, this malignant disease is primarily categorized into two distinct forms: the more prevalent non-small cell lung cancer (NSCLC), which constitutes about 85% of total cases, and the less common small cell lung cancer (SCLC) [1]. Among NSCLC cases, adenocarcinoma (LUAD) emerges as the most prevalent histological subtype, comprising approximately 40% of NSCLC cases, whereas squamous cell carcinoma (LUSC) accounts for 25–30% [2]. These statistics highlight the significant clinical impact and epidemiological importance of lung cancer subtypes in global oncology.

TMEM147, a recently discovered 25 kDa transmembrane protein, exhibits dual localization at both the endoplasmic reticulum (ER) membrane and nuclear envelope. This multi-spanning membrane protein, characterized by seven complete transmembrane domains, orients with its amino terminus projecting into the ER lumen and carboxyl terminus extending toward the cytosol [3,4]. The ER membrane, where TMEM147 predominantly resides, serves as a crucial site for essential cellular functions, particularly lipid biosynthesis [5]. Emerging evidence indicates that TMEM147 demonstrates widespread tissue distribution, implying its involvement in diverse physiological processes across multiple organ systems [6]. Recent investigations have revealed the clinical significance of TMEM147 in various pathological conditions, particularly in oncological contexts. Previous studies have identified this protein as significantly upregulated in colorectal cancer tissues relative to their normal counterparts, indicating its possible diagnostic value [7]. Additionally, research has revealed that TMEM147-AS1, an antisense long non-coding RNA derived from the same genomic locus, plays a crucial role in modulating tumor cell growth and metastasis in prostate cancer [8] . While these findings highlight the oncogenic relevance of the TMEM147 axis in certain malignancies, its functional role in LUAD remains unexplored. Our current study provides novel insights into the critical involvement of TMEM147 in LUAD progression and its potential prognostic value.

Here, we demonstrate for the first time that TMEM147 is significantly upregulated in LUAD tissues and functionally promotes cancer progression by stimulating cellular proliferation, migration, and invasion, as confirmed by multi-omics analyses and experimental validation. Through bioinformatics analysis, we identified potential transcription factors regulating TMEM147 expression. Furthermore, our investigation revealed significant correlations between TMEM147 expression patterns and clinical outcomes in LUAD patients. To elucidate the molecular mechanisms, we performed functional enrichment analyses and constructed protein interaction networks, while also examining the relationship between TMEM147 and tumor immune microenvironment characteristics. Gene ontology and pathway analyses were conducted to characterize the biological processes associated with TMEM147-regulated genes. Notably, our data suggest a possible link between TMEM147 levels and tumor-infiltrating immune cells in lung adenocarcinoma development.

## 2. Material and Methods

### 2.1 Data acquisition

Transcriptomic and clinical data for 33 cancer types, including lung adenocarcinoma (LUAD), were acquired from The Cancer Genome Atlas (TCGA). The LUAD cohort consisted of 535 tumor samples and 59 adjacent normal tissues. Furthermore, gene expression datasets (GSE102287 and GSE118370) were retrieved from the GEO database [9] .

### 2.2 Differentially expressed genes (DEGs) analysis

We employed bioinformatics approaches to detect differentially expressed genes by analyzing HTSeq-counts datasets from patient groups stratified by TMEM147 expression. The DESeq2 software (version 1.26.0) in R was utilized for statistical evaluation, with gene expression differences determined through Wilcoxon rank-sum testing. Transcripts meeting the threshold criteria of absolute log2 fold change >1 along with false discovery rate-adjusted P-values < 0.05 were classified as significant [10].

### 2.3 Gene set enrichment analysis

Functional enrichment analysis was conducted to investigate the biological roles of TMEM147-associated hub genes and differentially expressed genes. The “ClusterProfiler” R package was employed for Gene Ontology (GO) annotation and Kyoto Encyclopedia of Genes and Genomes (KEGG) pathway examination. Result visualization was achieved using the “ggplot2” graphical package. Additionally, pathway enrichment studies of TMEM147-related signaling cascades in LUAD were performed through Gene Set Enrichment Analysis (GSEA) coupled with the clusterProfiler toolkit [11,12].

### 2.4 Identification of 10 hub genes co-expressed with TMEM147

To investigate potential functional associations, we generated a protein-protein interaction (PPI) network using the STRING database for genes co-expressed with TMEM147 [13]. Subsequently, we identified 10 core hub genes from this network and validated their expression correlation with TMEM147 using the GEPIA platform. Furthermore, comparative analysis was performed to examine the differential expression patterns of these hub genes between normal lung tissues and lung adenocarcinoma (LUAD) samples.

### 2.5 Tumor Immune Infiltration Analysis

To quantify tumor microenvironment composition, we applied the ESTIMATE algorithm to calculate stromal and immune scores, reflecting respective cellular infiltration levels. Subsequently, single-sample gene set enrichment analysis (ssGSEA) was implemented through the GSVA R package to evaluate the abundance of 22 distinct immune cell populations. This approach enabled comprehensive characterization of tumor-infiltrating immune and stromal components [14].

To assess immune cell infiltration patterns, we calculated enrichment scores for 24 immune cell subtypes in tumors with high versus low TMEM147 expression. This analysis employed established immunocyte signature gene sets and individual tumor transcriptomic profiles to enable comparative evaluation between the two groups.

### 2.6 Cell culture

The human lung epithelial cell line BEAS2B and four LUAD-derived cell lines (A549, H1299, HCC827, and NCI-H1975) were acquired from ATCC. BEAS2B and A549 cells were cultured in Ham’s F-12K medium (Basal Media, L450KJ), while H1299, HCC827, NCI-H1975, and PC9 cell lines were grown in RPMI-1640 (Basal Media, L220KJ). Both culture media contained 10% fetal bovine serum (YeaSen, 40130ES76) and 1% penicillin-streptomycin (Basal Media, S110jv). All cell lines were kept at 37°C in a humidified atmosphere with 5% CO₂.

### 2.7 Western blotting

For immunoblotting analysis, tissue and cellular protein extracts were resolved by SDS-PAGE and electrotransferred to nitrocellulose membranes (Invitrogen). After blocking with 5% non-fat milk, membranes were probed overnight at 4°C with anti-TMEM147 primary antibody (1:1000 dilution; ABclonal, A17076). β-Tubulin (1:2000; Proteintech) served as an internal control for protein loading. After thorough washing, the membranes were probed with IRDye 800CW-labeled secondary antibodies (LI-COR; 1:20,000 dilution) for 90 minutes under ambient temperature conditions. Subsequent protein band visualization and quantitative analysis were performed using the Odyssey® Infrared Imaging System (LI-COR).

### 2.8 Transwell migration and invasion assay

The migratory and invasive potential of cells was evaluated using Transwell® assays (Corning, NY, USA), employing both uncoated and Matrigel-coated (BD Biosciences) polycarbonate membranes to distinguish between these two biological processes. Briefly, cells were seeded in the upper compartment containing serum-free medium, while complete growth medium was placed in the lower chamber. After a 48-hour incubation period, stationary cells adhering to the upper chamber membrane were carefully eliminated. Migrated/invaded cells attached to the lower membrane surface were subsequently fixed using methanol, subjected to crystal violet staining, and enumerated through microscopic analysis.

### 2.9 Wound-healing assay

A wound healing assay was performed to assess cellular migratory ability. Briefly, stable transfectants were seeded in 6-well plates (3×10^5 cells/well) and cultured to 100% confluence. The cell monolayer was then mechanically scratched using a sterile pipette tip (200 μL), followed by three PBS washes to remove detached cells. Subsequently, the wounded monolayers were incubated in medium lacking serum supplementation. Wound closure was monitored by capturing images at baseline (0 h) and 24 hours post-scratching to quantify migration rates.

### 2.10 Bioinformatic analysis of putative transcriptional regulators

To elucidate the transcriptional regulation of TMEM147, we employed a comprehensive bioinformatics approach integrating predictions from six independent databases: HTFtarget, JASPAR, ChIP_Atlas, FIMO_JASPAR, GTRD, and KnockTF. These resources were systematically interrogated to identify candidate transcription factors (TFs) potentially regulating TMEM147 expression. Intersection analysis of the prediction results from all six platforms was performed to determine consensus TFs with high confidence. The overlapping TF candidates were subsequently visualized using a Venn diagram to highlight the most promising regulatory factors.

### 2.11 Statistical analysis

Statistical computations and figure generation were executed in R (v4.2.1; R Foundation), employing the ggplot2 visualization package (v3.4.4). Differential expression of TMEM147 was assessed by a nonparametric test: Wilcoxon rank-sum test for independent samples. The predictive accuracy was assessed through ROC analysis conducted with the pROC package (version 1.17.0.1), with area under the curve (AUC) values between 0.5 (random chance) and 1.0 (perfect discrimination) quantifying predictive accuracy.

For prognostic assessment, we employed Kaplan-Meier survival curves with log-rank testing, complemented by both univariate and multivariate Cox regression modeling. Associations with clinicopathological parameters were examined using Pearson’s χ² test. To ensure methodological robustness, key findings were independently validated using GraphPad Prism software (version 10.0).*, P < 0.05; **, P < 0.01; ***, P < 0.001.

## 3 Results

### 3.1 TMEM147 is highly expressed in lung adenocarcinoma and is associated with poor prognosis

Our initial investigation analyzed TMEM147 expression patterns across various cancers using TCGA datasets. The results demonstrated significant upregulation of this gene in 33 cancer types, particularly in lung adenocarcinoma (Figure.1A). Further analysis of TMEM147 expression in matched tumor and adjacent normal tissues across 23 cancer types confirmed its significant upregulation in tumors (Figure.1B). Meanwhile, Analysis of the TCGA LUAD cohort revealed significantly upregulated TMEM147 expression in tumor tissues compared with paired normal controls (Figure.1C-D). This finding was further validated in two independent GEO datasets (GSE102287 and GSE118370), where TMEM147 consistently showed elevated expression levels in LUAD specimens (Figure.1E-F). To investigate the clinical relevance of TMEM147 overexpression, we conducted survival analyses using three distinct prognostic parameters: overall survival (OS), disease-specific survival (DSS), and progression-free interval (PFI). Kaplan-Meier curves indicated that high TMEM147 expression was significantly associated with poorer clinical outcomes across all evaluated endpoints (Figure. 1G-I), Furthermore, we constructed a receiver operating characteristic (ROC) curve to evaluate the diagnostic potential of TMEM147. The area under The curve (AUC) value was 0.831, indicating a strong prognostic value for LUAD detection (Figure.1J), suggesting its potential role as an adverse prognostic marker in LUAD.

**Fig. 1.**
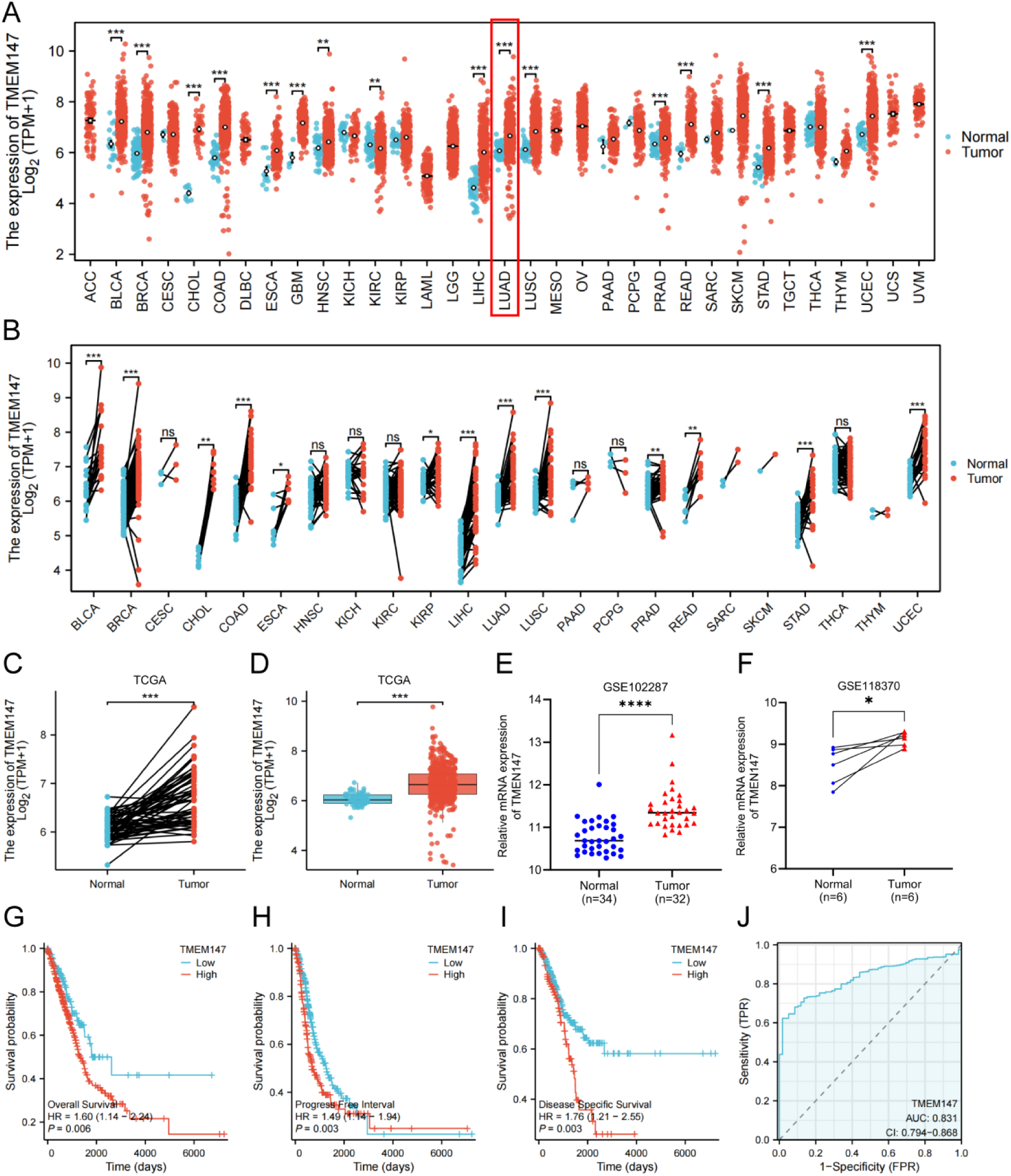
The expression and survival prognosis of TMEM147 in LUAD datasets in TCGA and GEO. (A) Comparative analysis of TMEM147 transcript levels between tumor and adjacent normal tissues across 23 cancer types from TCGA. (B) Paired-sample evaluation of TMEM147 dysregulation in matched tumor-normal specimens. (C) Differential expression profiling in LUAD versus normal lung tissues. (D) Validation of TMEM147 overexpression in tumor samples compared with their paired normal counterparts. (E-F) Independent confirmation of elevated TMEM147 expression in LUAD datasets from GEO. (G-I) Kaplan-Meier survival analysis demonstrating significant differences in OS, DSS and PFI between LUAD patients stratified by TMEM147 expression. (J) Diagnostic performance assessment through ROC curve analysis evaluating TMEM147’s ability to discriminate LUAD tissues. *p < 0.05, **p < 0.01, ***p < 0.001, NS, no significance.

### 3.2 Correlation between TMEM147 expression level and clinical features

We extracted clinical records of LUAD cases from the TCGA database to investigate associations between TMEM147 expression and patient characteristics. After applying exclusion criteria for incomplete records, the final cohort comprised 539 patients with a median age of 61.5 years (IQR: 49.25-70.00), including 46.4% male participants (Supplementary Table 1). Using Kruskal-Wallis tests on TMEM147 expression-stratified groups, we identified significant correlations (p<0.05) with multiple clinicopathological parameters: demographic factors (age, gender, race), disease characteristics (TNM stage, histological grade, tumor location), progression markers (pathological stage, tumor status), treatment outcomes (residual tumor), and behavioral factors (smoking history) (Figure.2A-O). TMEM147 expression showed particularly strong associations with OS and DSS, though no significant relationship was observed with PFI (Figure.2P).

### 3.3 FLI1 is a potential regulator of TMEM147 in LUAD

To delineate the transcriptional regulatory network controlling TMEM147 overexpression, we employed bioinformatic approaches to predict potential upstream regulatory factors. Integration of six prediction databases identified FLI1 as a candidate transcription factor (Figure.3A). Subsequent differential expression analysis revealed that FLI1 levels were markedly reduced in LUAD samples from both the TCGA cohort and the GSE102287 dataset (Figure.3B-C). Notably, TMEM147 exhibited a significant inverse correlation with FLI1 expression (Figure.3D), suggesting a potential regulatory relationship. The motif map illustrates transcription factor binding sequences, and is predicted by JASPAR database. Those with a score greater than 10 are selected for plotting, and the relative positions of the predicted binding site sequences are shown in the figure (Figure.3F-F).These results support a potential role for FLI1 as a regulator of TMEM147.

### 3.4 Functional analysis of TMEM147 in LUAD

To investigate TMEM147-related molecular features in LUAD, cases were dichotomized into high- and low-expression subgroups based on TMEM147 transcript quantification. Gene expression profiling was subsequently conducted using DESeq2, applying stringent criteria (adjusted p-value <0.05 and absolute log2 fold-change >1) to identify statistically significant differentially expressed genes. This approach identified 1,272 significantly dysregulated genes, comprising 646 upregulated and 626 downregulated transcripts in the high-expression group compared to controls (Figure.4A; Supplementary Table 2). Subsequent HTSeq-Counts evaluation revealed the 10 most prominently altered genes, ranked by expression magnitude (Figure. 4B).Finally, GO enrichment analysis of differential genes showed that co-expressed genes were mainly involved in ribosomal protein biogenesis, mitochondrial gene expression and oxidative phosphorylation (BP). And the inner mitochondrial membrane, ribosome (CC); Ribosome structure composition, Redox activity (MF) and other related functions. KEGG pathway analysis also found that TMEM147 is enriched in several signaling pathways associated with ribosome and oxidative phosphorylation.These results provide a more comprehensive perspective for understanding the molecular mechanism of lung cancer.

**Fig. 2.**
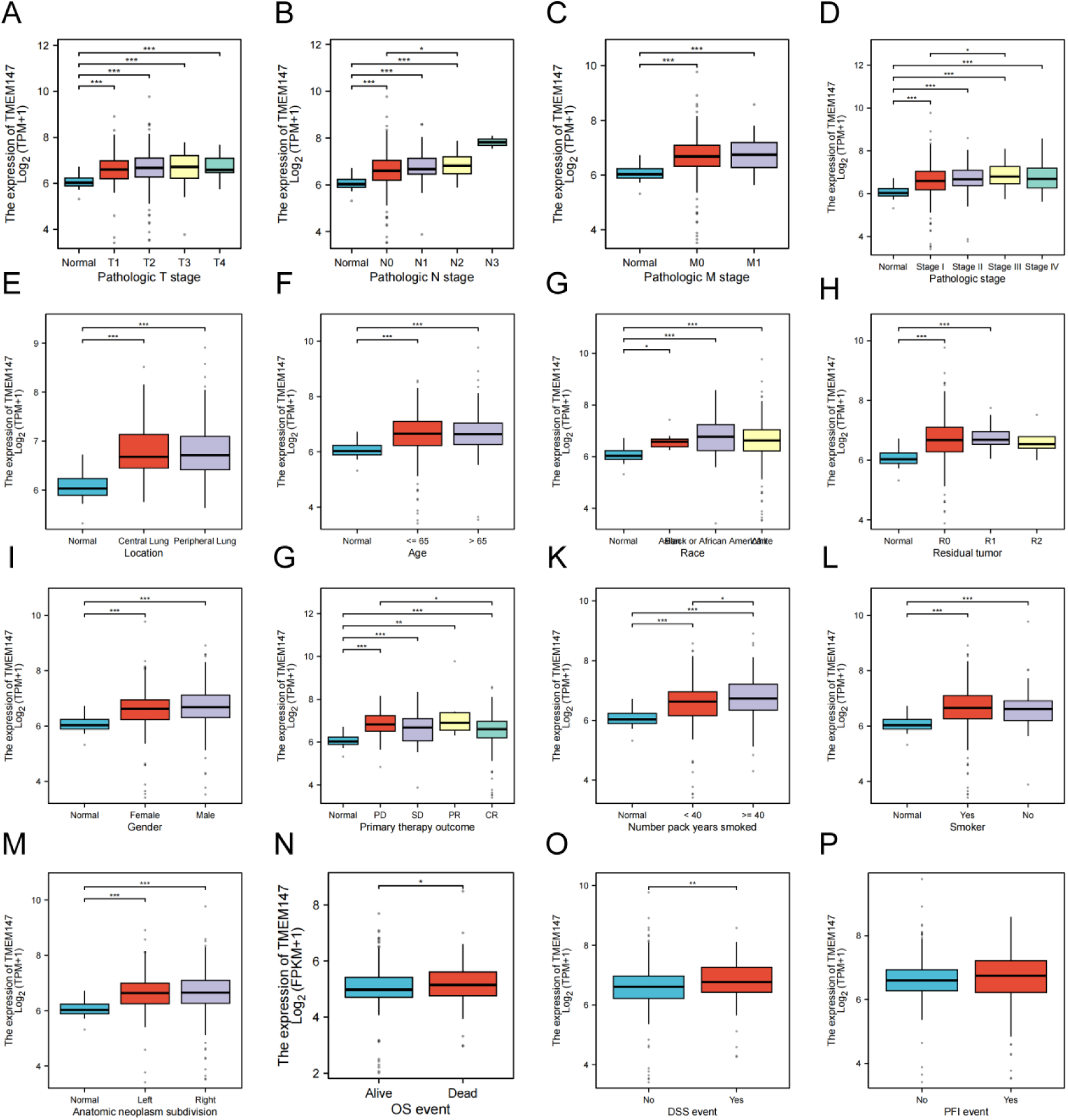
Correlation of TMEM147 expression with clinicopathological characteristics. (A) T stage. (B) N stage. (C) M stage. (D) Pathological stage. (E) Location. (F) Age. (G) Race. (H) Residual tumor. (I) Gender. (J) Primary therapy outcome. (K) Number pack years smoked. (L) Smoker. (M) Anatomic neoplasm subdivision. (N) OS event. (O) DSS event. (P) PFI event. *p<0.05,**p<0.01, ***p<0.001

**Fig. 3.**
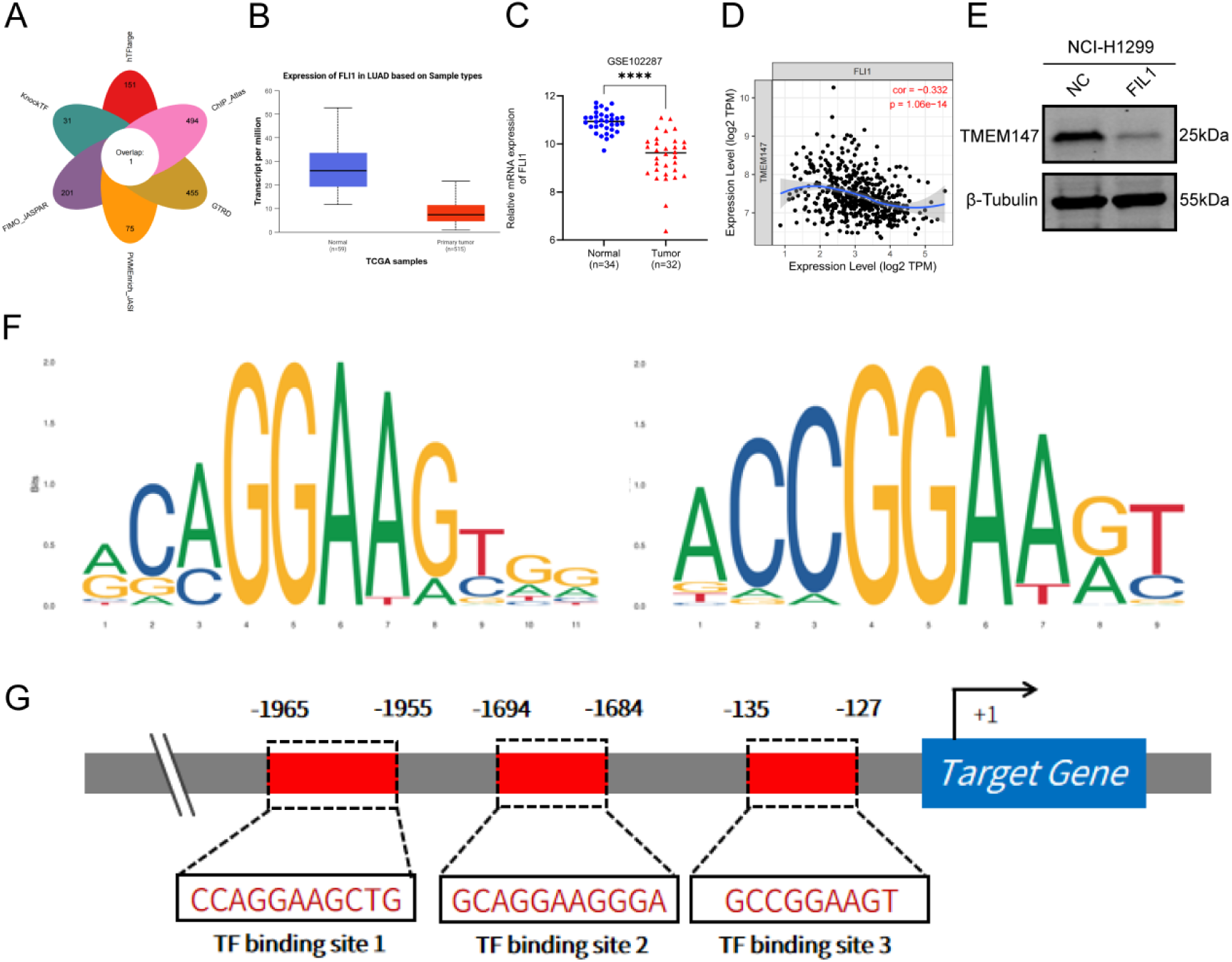
Transcription Factor Prediction. (A) A single transcriptional regulator was identified using multiple prediction tools (n=6). (B-C) The expression levels of transcription factors were verified in TCGA and GSE102287 LUAD datasets. (D) Correlation analysis between FLI1 and TMEM147. (E) Motift map of transcription factor binding sequence. (F) Predicted binding sites of FLI1 on the TMEM147 promoter region of transcription factor binding site sequences.

**Fig. 4.**
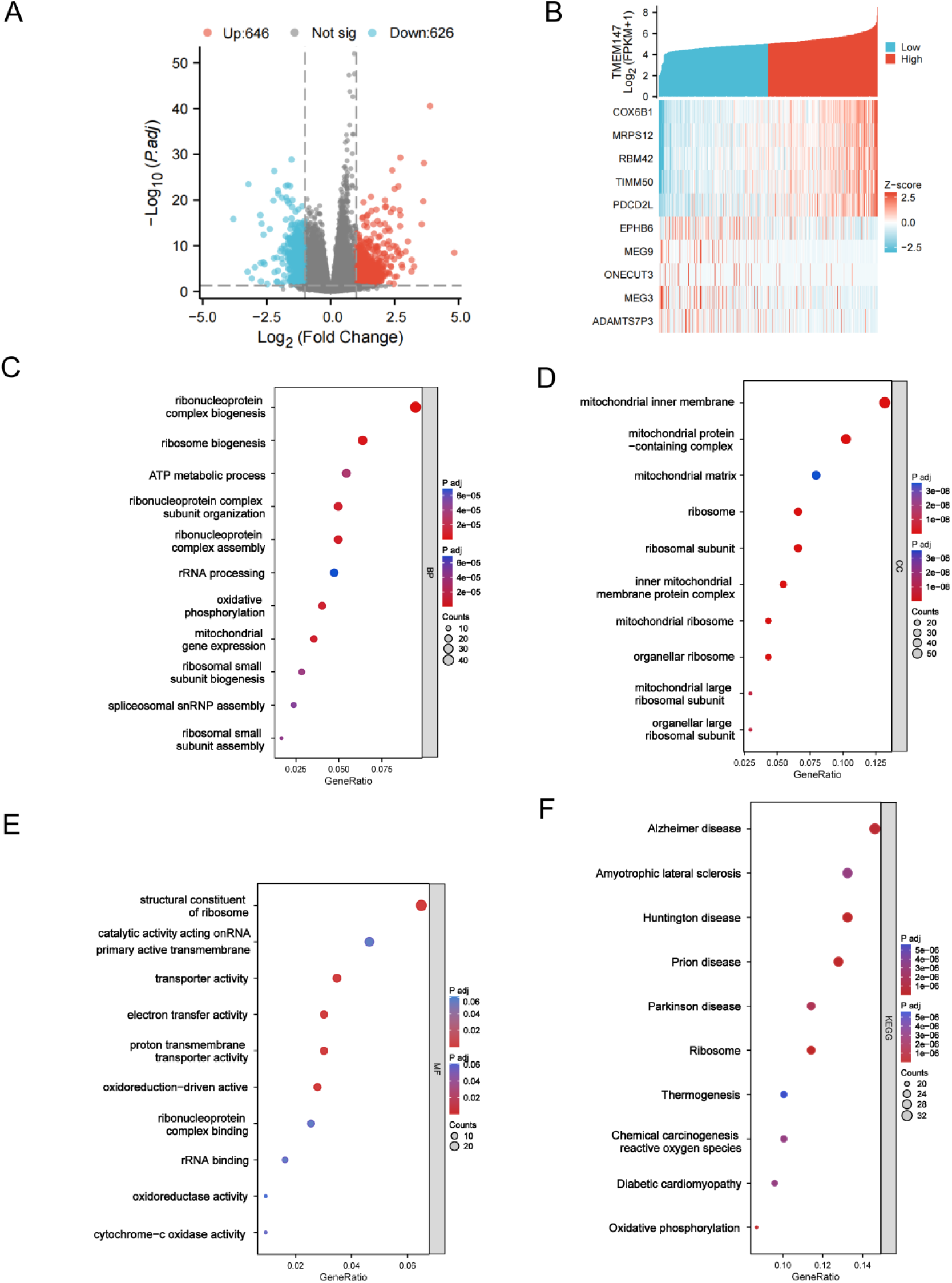
Functional enrichment analyses of TMEM147-relatedgenesin LUAD. (A-B) Volcano plots of the DEGs and heat maps showing the top10DEGs.(C) BP analysis. (D) CC analysis. (E) MF analysis. (F) KEGG Pathway analysis.

### 3.5 Relationship between TMEM147 expression and immune cell infiltration

Using single-sample gene set enrichment analysis (ssGSEA), we systematically evaluated and measured the association of TMEM147 expression patterns with immune cell infiltration in the tumor microenvironment (Figure.5A). We further confirmed that TMEM147 expression was negatively correlated with Tem, Tcm, Eosinophils, and T helper cells and positively correlated with Tgd cells and Th2 cells (Figure. 5B-G). Meanwhile, Analysis of the somatic cell number alteration (SCNA) module through the TIMER database revealed significant correlations between TMEM147 copy number variations (deletion or amplification) and immune cell infiltration levels in LUAD (Figure.5H). To further investigate TMEM147’s immunomodulatory role, we stratified LUAD patients into high- and low-expression groups according to TMEM147 transcript levels. This classification enabled comparative evaluation of tumor immune microenvironment features between the subgroups (Figure.5I). We found that Tgd and Th2 cells increased (P < 0.05), and Tem, Tcm, Eosinophils, and T helper cells decreased (P < 0.05) in the TMEM147 high expression group. These results confirm that elevated TMEM147 expression in LUAD is closely related to immune cellsuppression.

**Fig. 5.**
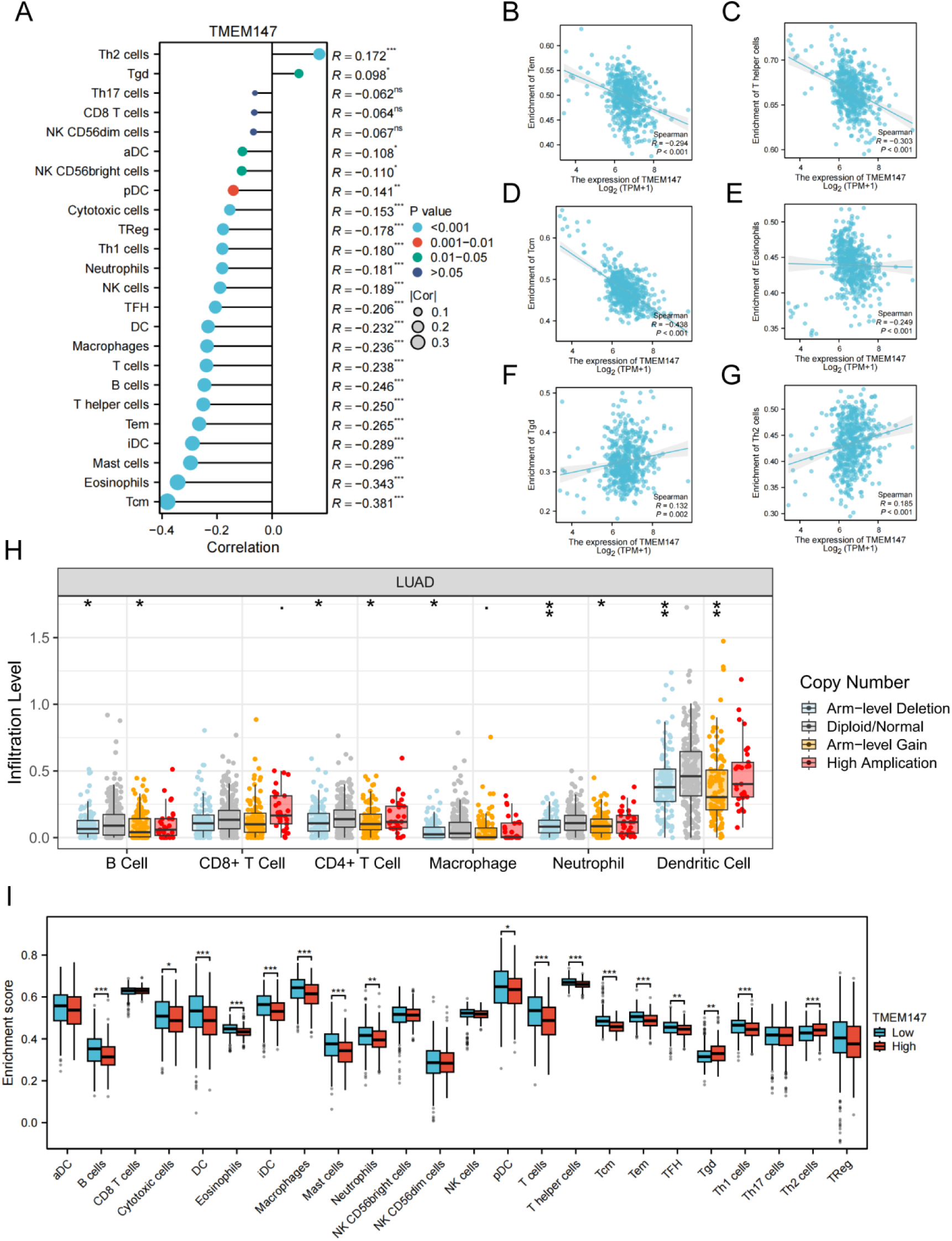
The immune infiltration of TMEM147 expression in LUAD. (A) Forest plot illustrating associations between TMEM147 transcript abundance and infiltration levels of 24 immune cell subtypes. (B-G) Comparative scatter plots demonstrating differential infiltration patterns of Tgd, Th2, Tem, central memory Tcm, eosinophils, and T helper cells across TMEM147 expression groups. (H) Somatic copy number alteration (SCNA) analysis revealing significant correlations between TMEM147 genomic alterations and immune infiltration. (I) Comprehensive correlation matrix evaluating associations between all 24 immune cell types and TMEM147 expression. *p < 0.05, **p < 0.01, ***p < 0.001, NS, no significance.

**Fig. 6.**
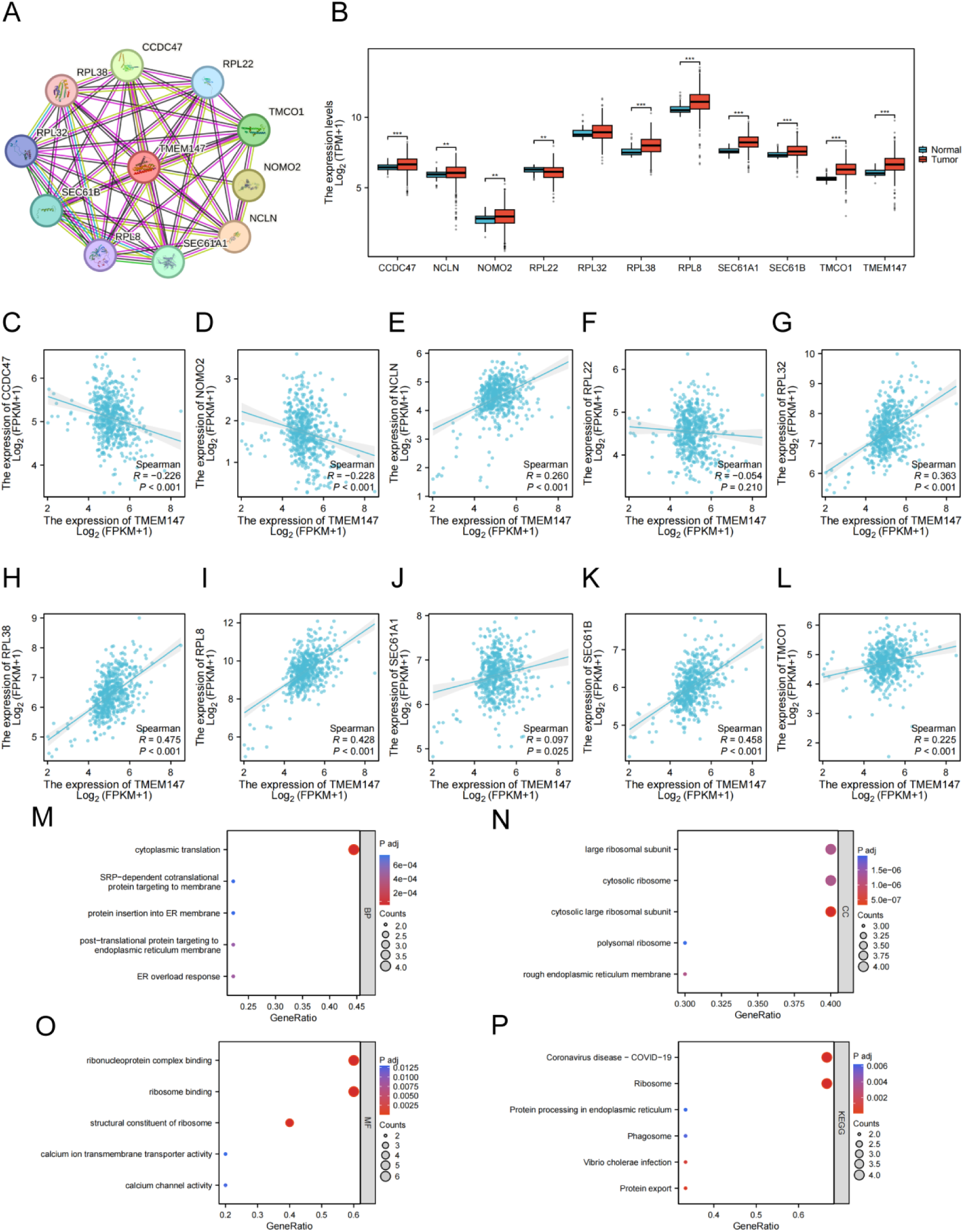
Protein-protein interaction network and function enrichment in LUAD. (A) Protein-protein interaction network of TMEM147; (B) Expression profiles of 10 genes functionally associated with TMEM147 in lung adenocarcinoma from TCGA; (C-L) Correlation heatmaps demonstrating co-expression patterns between TMEM147 and its associated genes; (M-P) Functional enrichment analysis showed that there were significant pathways of GO and KEGG enrichment.

**Fig. 7.**
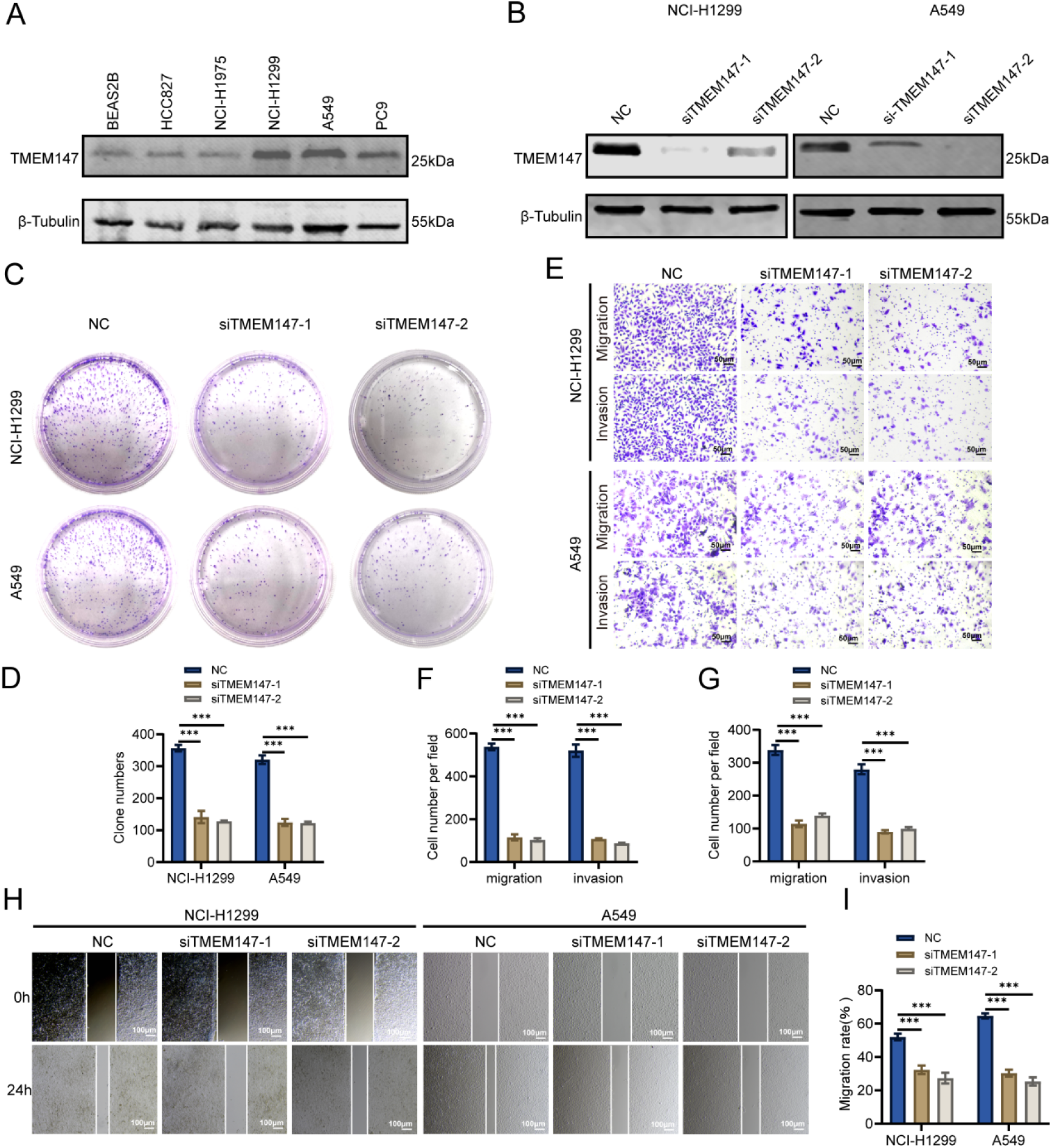
TMEM147 promotes the proliferation, migration and invasion of LUAD in vitro. (A)The protein expression levels of TMEM147 in BEAS2B, HCC827, NCI-H1975, NCI-H1299, A549 and PC9 cell lines were detected. (B) Western blot analysis of TMEM147 expression in NCI-H1299 and A549 cells (siTMEM147-1, siTMEM147-2 and control). (C-D) NCI-H1299 and A549 cells were used to determine clone formation assays. (E-G) Transwell assays showing migration and invasion abilities. (H-I) Wound-healing assays. Scale bars: Scale bars: 100 μm (100×), 50 μm (200×).*p < 0.05, **p < 0.01, ***p < 0.001, NS, no significance.

### 3.6 Identification of 10 hub genes co-expressed with TMEM147

Through STRING analysis, we detected 10 coregulated genes interacting with TMEM147. including CCDC47, NCLN, NOMO2, RPL22, RPL32, RPL38, RPL8, SEC61A1, SEC61B and TMCO1 (Figure.6A). Next, We subsequently quantified these 10 candidate genes in LUAD specimens, revealing consistently elevated expression across all identified interactors. (Figure.6B). To characterize the molecular network involving TMEM147 in lung adenocarcinoma, we analyzed the co-expression patterns of this gene with ten functionally associated targets. Our results demonstrated significant positive correlations (p<0.05) between TMEM147 transcript levels and eight ribosomal/secretory pathway genes, including NCLN, multiple RPL family members (RPL22, RPL32, RPL38, RPL8), SEC61 subunits (SEC61A1, SEC61B), and TMCO1 (Figure.6C-L). To elucidate the biological roles of TMEM147-associated genes in LUAD, GO and KEGG pathway analyses were conducted. (Figure.6M-P).

### 3.7 TMEM147 promotes the proliferation, migration and invasion of lung adenocarcinoma

To further investigate the function of TMEM147 in lung adenocarcinoma, we validated the expression of TMEM147 in lung epithelial cells and lung adenocarcinoma cell lines (Figure.7A). Subsequently, we used siRNA to knock down TMEM147 expression in NCI-H1299 and A549 cells, which exhibit relatively high TMEM147 expression.Western blot analysis confirmed the efficiency of TMEM147 knockdown (Figure.7B). Clonogenic assays demonstrated that TMEM147 silencing markedly reduced proliferative capacity in both NCI-H1299 and A549 cell lines (Figure.7C-D). Transwell assay showed that TMEM147 knockdown significantly reduced the migration and invasion abilities of NCI-H1299 and A549 cells (Figure.7E-G).Wound-healing assays further confirmed that TMEM147 inhibition inhibited the migration of NCI-H1299 and A549 cells (Figure.7H-I).

## 4. Discussion

Despite recent advances in radiation, systemic, molecularly-targeted and immune-based treatments, Patients with advanced NSCLC continue to demonstrate unfavorable clinical outcomes, exhibiting consistently suboptimal 5-year survival probabilities [15]. In this study, we identified TMEM147 as a potent oncogene for LUAD proliferation and metastasis, and systematically revealed the oncogenic role of transmembrane protein TMEM147 in lung adenocarcinoma (LUAD) and its regulatory mechanism on immunosuppressive tumor microenvironment (TME) for the first time.Through combined multi-omics and experimental validation, we demonstrated that elevated TMEM147 expression correlates with adverse clinical outcomes in LUAD and plays a central role in enhancing tumor cell proliferation, motility, and invasiveness. The following is an in-depth discussion of the research results from multiple dimensions.

Data set analysis showed that TMEM147 expression was significantly increased in lung adenocarcinoma tissues compared with normal controls. and its high expression was closely related to TNM stage, pathological grade and worse patient survival. ROC curve analysis showed that TMEM147 had a high diagnostic efficiency (AUC=0.831), indicating its potential utility as a standalone prognostic indicator.This finding is consistent with previous studies in colorectal cancer [7], but the present study is the first to validate its clinical value in LUAD. Notably, TMEM147 expression was significantly correlated with clinical parameters such as smoking history and tumor residual status, implying its possible involvement in the malignant transformation process driven by environmental carcinogens such as tobacco exposure.

Subsequently, FLI1 was predicted and validated as a key transcriptional regulator of TMEM147 by joint analysis of six data libraries. As a member of the ETS family, FLI1 has a dual role in promoting or suppressing cancer in a variety of cancers [16].Interestingly, FLI1 expression was down-regulated in LUAD (Figure. 3B-C) and showed a significant negative correlation with TMEM147, suggesting that it may function as a tumor suppressor by inhibiting TMEM147 transcription. This finding broadened our understanding of the epigenetic regulatory network of LUAD.However, the mechanisms of how FLI1 specifically regulates LUAD via TMEM147, such as DNA methylation or chromatin remodeling, still need to be further verified by ChIP-seq and promoter activity experiments.

Functional enrichment analysis showed that TMEM147-related differential genes were significantly enriched in ribosomal biosynthesis and oxidative phosphorylation pathways. Combined with PPI network analysis, TMEM147 is co-expressed with multiple ribosomal proteins (e.g., RPL22, RPL8) and endoplasmic reticulum transporters (e.g., SEC61A1),This suggests that it may be through ER-ribosome coupling: TMEM147 is localized in the endoplasmic reticulum membrane and may mediate the co-translation and transport of nascent peptide chains through SEC61A1 (an endoplasmic reticulum transport channel protein) to ensure the dynamic coupling of ribosomes and endoplasmic reticulum [5]. Ribosome quality control: Ribosomal proteins such as RPL22 are involved in ribosome assembly and rRNA modification, and TMEM147 may affect ribosome maturation by regulating its stability [17]. KEGG also showed that TMEM147-related genes were enriched in the OXPHOS pathway, and the mechanism may be involved in: Regulation of mitochondrial-endoplasmic reticulum contacts (MERCs) : TMEM147 is localized to the endoplasmic reticulum membranes and may regulate Ca^2+^ signaling (affecting mitochondrial ATP synthesis) or lipid transport (such as cardiolipin synthesis) through MERCs [18] .Ribosome-oxphos synergy: ribosomal biosynthesis requires a large amount of ATP (provided by OXPHOS), and TMEM147 may indirectly promote OXPHOS demand by enhancing ribosomal activity [19].Regulation of electron transport chain (ETC) components: SEC61A1 is involved in the co-translational insertion of mitochondrial membrane proteins (such as complex IV subunits), and TMEM147 may affect ETC function by regulating transport efficiency [20].

In addition, the present study revealed an association between TMEM147 expression and the pattern of tumor immune infiltration. Th2 cell and Tgd cell infiltration was increased in the high TMEM147 group, while effector memory T cells (Tem) and helper T cells were decreased. Th2 polarization usually promotes the differentiation of M2-type macrophages by secreting IL-4/IL-13 [21], while Tgd cells have immunosuppressive properties in a variety of cancers [22], and the two together constitute a tumor-promoting immune microenvironment. Meanwhile, TMEM147 copy number variation (SCNA) was significantly correlated with the level of immune cell infiltration, suggesting that it may indirectly regulate TME through genomic instability. These findings provide evidence for the immunotherapy potential of TMEM147. In the future, the combination of TMEM147 inhibitors and immune checkpoint blockade (such as anti-PD-1) or the development of interventions targeting Th2/Tgd cells can be explored.

Of note, the metabolic heterogeneity of NSCLC has been shown to significantly influence treatment response. Studies have shown that different cell subsets within tumors can drive treatment resistance through differential utilization of nutrient substrates (e.g., glucose, glutamine) or activation of alternative metabolic pathways (e.g., oxidative phosphorylation, fatty acid oxidation) [23]. In this study, we found that TMEM147 promotes LUAD progression by regulating ribosome biosynthesis and oxidative phosphorylation (OXPHOS), suggesting that it may be a key regulator of metabolic heterogeneity. Of particular interest, TMEM147-mediated OXPHOS activation may enhance cancer cell survival under targeted therapy or chemotherapy-highly consistent with a recently identified metabolic adaptive resistance mechanism [24].

The identified roles of TMEM147 in immunosuppression and metabolic reprogramming align with several emerging therapeutic strategies currently under clinical evaluation. Notably, interventions targeting TMEM147-associated pathways may overcome resistance to existing therapies: Among the OXPHOS inhibitors, IACS-010759 (NCT03291938) showed potential in advanced solid tumors (including NSCLC) dependent on mitochondrial metabolism, especially for TMEM147-driven OXPHOS isoforms [25]. Immunomodulatory strategies focused on reversing TMEM147-mediated immunosuppression: anti-IL-4 /IL-13 antibody Dupilumab combined with PD-1 inhibitor (NCT05069935) antagonized Th2 polarization [26]. Ribosome biosynthesis inhibitor CX-5461 (NCT02719977) is the first RNA polymerase I inhibitor targeting the ribosome synthesis pathway in advanced solid tumors, and its early efficacy against Myc-driven tumors suggests potential applicability to TMEM147-overexpressing LUAD [27]. These advances provide a clinical translation framework for precision treatment based on TMEM147 molecular typing.

Beyond its role in immune modulation and ribosomal functions, emerging evidence implicates dysregulated Hippo-YAP/TEAD signaling as a critical mediator of therapy resistance in lung cancer.The transcriptional co-activator YAP, upon Hippo pathway inactivation, complexes with TEAD to drive pro-survival and pro-proliferative genes, conferring resistance to targeted therapies and conventional treatments in NSCLC [28–29]. Notably, several YAP/TEAD inhibitors are under clinical investigation to overcome this resistance [30]. While our study did not directly explore TMEM147’s interaction with Hippo signaling, the observed TMEM147-driven enrichment in ribosomal biogenesis and OXPHOS pathways may indirectly sustain YAP/TEAD activity. Ribosomal hyperactivation and mitochondrial energetics are known to support oncogenic transcription factors like YAP [31]. Future studies should examine whether TMEM147 promotes therapy resistance via YAP/TEAD axis potentiation, potentially positioning TMEM147 as a co-target with novel YAP/TEAD inhibitors.

## Conclusions

TMEM147 is upregulated in LUAD, correlating with poor prognosis (AUC=0.831) and advanced clinicopathology. FLI1 inversely regulates TMEM147, linked to ribosome biogenesis and OXPHOS. TMEM147 promotes immunosuppression via Th2/Tgd cell infiltration and Tem reduction. Silencing TMEM147 inhibits LUAD proliferation, migration, and invasion. These findings highlight TMEM147 as a prognostic biomarker and therapeutic target, warranting exploration of FLI1-mediated regulation and immunotherapeutic synergy.

## Data availability

Data will be made available on request.

## Data Availability

All relevant data are within the manuscript and its Supporting Information files

## Acknowledgments

Not applicable

